# A randomized placebo-controlled clinical trial of Nicotinamide Riboside and Pterostilbene supplementation in experimental muscle injury in elderly subjects

**DOI:** 10.1101/2021.10.04.21264504

**Authors:** Jonas Brorson Jensen, Ole L. Dollerup, Andreas B. Møller, Tine B. Billeskov, Emilie Dalbram, Sabina Chubanava, Ryan W. Dellinger, Kajetan Trost, Thomas Moritz, Steffen Ringgaard, Niels Møller, Jonas T. Treebak, Jean Farup, Niels Jessen

## Abstract

**Background:** Maintenance and regeneration of functional skeletal muscle are dependent on a sufficient pool of muscle stem cells (MuSCs). During ageing there is a functional decline in this cellular pool which influences the regenerative capacity of skeletal muscle. Preclinical evidence have suggested that Nicotinamide Riboside (NR) and Pterostilbene (PT) can improve muscle regeneration e.g. by increasing MuSC function. The objective of the present study was to investigate if NRPT supplementation promotes skeletal muscle regeneration after muscle injury in elderly humans by improved recruitment of MuSCs.

**Methods:** In a randomized, double-blinded, placebo-controlled trial, 32 elderly men and women (55-80 yr) received daily supplementation with either NRPT (1000 mg NR + 200 mg PT) or matched placebo. Two weeks after initiation of supplementation, a skeletal muscle injury was applied in the vastus lateralis part of the quadriceps femoris muscle by electrically induced eccentric muscle work in a dynamometer. Skeletal muscle biopsies were obtained pre, 2h, 2, 8, and 30 days post injury. The main outcome of the study was change in MuSC content 8 days post injury.

**Results:** 31 enrolled subjects completed the entire protocol. The muscle work induced a substantial skeletal muscle injury in the study participants and was associated with release of myoglobin and creatine kinase, muscle soreness, tissue edema, and a decrease in muscle strength. MuSC content increased by 107% 8 days post injury (p= 0.0002) but with no effect of NRPT supplementation (p=0.58 for supplementation effect). MuSC proliferation and cell size revealed a large demand for recruitment post injury but was not affected by NRPT. Furthermore, histological analyses of muscle fiber area, internal nuclei and embryonic Myosin Heavy Chain showed no effect of NRPT supplementation.

**Conclusion:** Daily supplementation with 1000 mg NR + 200 mg PT is safe but does not improve recruitment of the MuSC pool or other measures of muscle recovery in response to injury or subsequent regeneration in elderly subjects.

## Introduction

Skeletal muscle mass is positively correlated with longevity and is inversely correlated with negative health outcomes in elderly subjects (1, 2). A sufficient and functional pool of muscle stem cells (MuSCs) is required to repair and maintain skeletal muscle (3). MuSCs mostly reside in a quiescent state (reversible cell cycle arrest) but is activated and proliferate upon environmental signals such as injury and other pathological conditions. The majority of proliferating MuSCs undergo differentiation and fusion into myofibers while a minor subset returns to quiescence and thereby maintains the MuSC pool. However, in rodent models of ageing, the MuSC content and function decline with the course of ageing and this is directly related to impaired myofiber regeneration (3).

Nicotinamide adenine dinucleotide (NAD^+^) homeostasis is critical for cell and organismal function. NAD^+^ and associated metabolites act as co-enzymes in redox reactions and co-substrates for other classes of enzymes including sirtuins and poly(adenosine diphosphate–ribose) polymerases (4). In humans, age and NAD^+^ levels are negatively correlated in several types of tissues (5-7). Thus, a decline in cellular NAD^+^ levels is a proposed hallmark of ageing. NAD^+^ synthesis is largely dependent on the salvage pathway in which nicotinamide phosphoribosyltransferase (NAMPT) is rate-limiting (8). Studies have revealed an age-associated decrease in NAMPT levels in certain tissues including skeletal muscle (9-11). This indicates that lower NAMPT levels are partly responsible for decreased NAD^+^ levels. In murine skeletal muscle, ablation of NAMPT causes substantial NAD^+^ lowering and progressive muscle degeneration (12-14). In these models, muscle function is partly rescued by Nicotinamide Riboside (NR) which bypasses NAMPT in generation of NAD^+^.

The transition of MuSCs from a quiescent state to cell cycle entry requires a shift in substrate metabolism, which is partly sirtuin-dependent (15). Thus, preclinical studies have shown beneficial effects of NAD^+^ increasing strategies in relation to MuSC quantity, function and overall enhancement of the muscle regeneration process (16, 17). Specifically, NR supplementation was shown to enhance regeneration after muscle injury in aged mice by SIRT1-mediated upregulating of genes related to oxidative phosphorylation in MuSCs (17). In addition, preclinical data suggest that muscle repair can be improved by addition of the polyphenol Pterostilbene (PT), a suggested activator of the sirtuin familiy (18). PT is an analog to Resveratrol but with higher bioavailiability due to the presence of two methoxy groups (19). Investigations of combined NR and PT effects in humans are limited but recent studies show beneficial effects of NRPT supplementation on functional strength parameters in both elderly individuals and patients with Amyotrophic Lateral Sclerosis (20, 21).

In the present study, we investigated if NRPT supplementation improves recruitment of the MuSC pool, and thereby skeletal muscle regeneration, after an experimentally induced injury in elderly humans in a randomized placebo-controlled design.

## Results

### Anthropometrics and activity level

Enrollment and data collection have been performed in the period from February 2019 to September 2020. 46 people were invited for screening of which 14 did not meet inclusion criteria (n=6) or declined to participate (n=8). 32 participants were enrolled and randomly assigned to supplementation with either NRPT (NR: 1000 mg/daily, PT: 200 mg/daily) or matched placebo for 45 days (14 days before and 30 days after muscle injury, Figure 1). 31 participants completed the study with one drop-out (placebo group) due to unacceptable pain related to the initial muscle biopsy. Anthropometric data of the two groups are presented in Table 1.

**Fig. 1:**
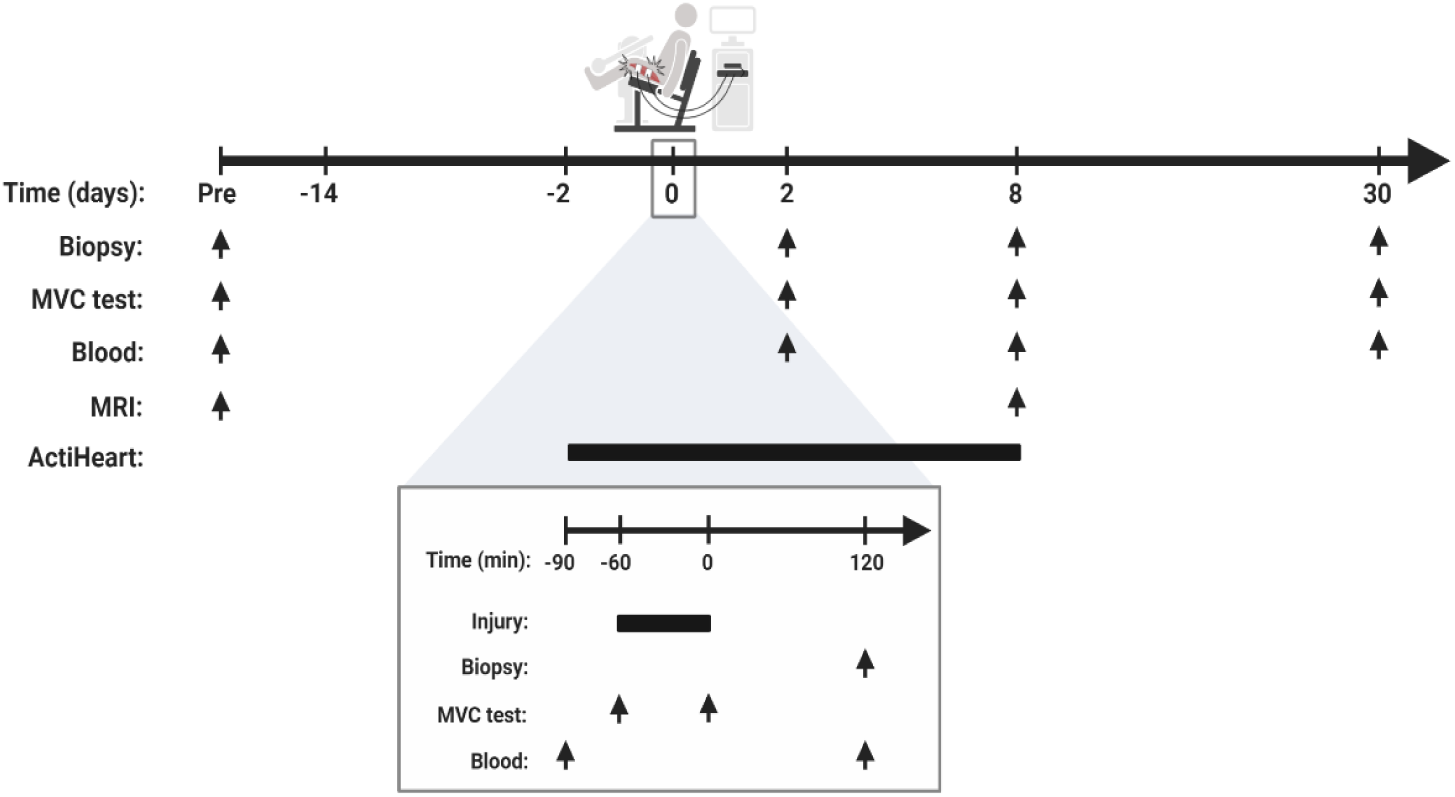
Study overview. Participants were examined before initiation of NRPT or matched placebo supplementation (Pre). Supplementation started 14 days prior to induction of skeletal muscle injury and continued until 30 days post injury. Skeletal muscle injury was induced at time point 0 by electrical induced contractions combined with eccentric work in a dynamometer (200 repetitions, 100 slow and 100 fast). The bottom line gives a detailed description of the injury day. The arrows indicate timing of skeletal muscle biopsy, MVC test, blood samples, and MR-imaging, respectively. Bars indicate a time span. Skeletal muscle biopsies were obtained from the uninjured leg prior to supplementation start and from the injured leg after induction of injury. MVC = maximal voluntary contraction.

**Tab. 1:** Participant characteristics. Data are expressed as mean ± SD. d/s = angle degree per second. MVC = maximal voluntary contraction. mA = milliampere. BPM = beats per minute. CPM = counts per minute. REE = resting energy expenditure. AEE = active energy expenditure. TEE = total energy expenditure.

|  |  | **PLA** | **NRPT** | **P-value** |
| --- | --- | --- | --- | --- |
| **n** |  | 15 | 16 |  |
| **Sex (n, male/female)** |  | 9/6 | 9/7 |  |
| **Injured leg (dom./non-dom.)** |  | 7/8 | 6/9 |  |
| **Age (years)** |  | 63 ± 6 | 69 ± 7 | 0.01 |
| **Height (cm)** |  | 174 ± 0 | 173 ± 0 | 0.86 |
| **Waist/Hip-ratio** |  | 1.0 ± 0.1 | 1.0 ± 0.1 | 0.65 |
| **Weight (kg)** | **Pre** | 75.8 ± 10.9 | 71.3 ± 8.2 | 0.21 |
|  | **Post** | 74.3 ± 9.6 | 71.0 ± 8.0 | 0.31 |
| **BMI (kg/m2)** | **Pre** | 25.0 ± 2.5 | 23.8 ± 2.3 | 0.16 |
|  | **Post** | 25.1 ± 2.3 | 23.7 ± 2.4 | 0.11 |
| **Lean mass (kg)** | **Pre** | 48.2 ± 8.5 | 45.5 ± 7.3 | 0.35 |
|  | **Post** | 45.9 ± 7.3 | 45.4 ± 7.3 | 0.86 |
| **Fat mass (kg)** | **Pre** | 25.8 ± 5.4 | 25.0 ± 5.8 | 0.69 |
|  | **Post** | 25.1 ± 5.6 | 24.8 ± 5.8 | 0.87 |
| **Fat (%)** | **Pre** | 33.8 ± 6.0 | 34.4 ± 7.6 | 0.82 |
|  | **Post** | 34.3 ± 6.5 | 34.2 ± 6.8 | 0.97 |
| **Compliance (%)** |  | 96 ± 5 | 96 ± 4 | 0.95 |
| **Work, 30 d/s (J/kg)** |  | 1128 ± 645 | 1124 ± 525 | 0.98 |
| **Work, 180 d/s (J/kg)** |  | 1115 ± 621 | 1121 ± 527 | 0.98 |
| **Work 30 d/s/(MVC)** |  | 6.6 ± 2.9 | 7.3 ± 2.7 | 0.49 |
| **Work 180 d/s/(MVC)** |  | 6.5 ± 2.6 | 7.2 ± 2.3 | 0.41 |
| **Current, 30 d/s (mA)** |  | 54 ± 13 | 60 ± 14 | 0.21 |
| **Current, 180 d/s (mA)** |  | 65 ± 17 | 69 ± 15 | 0.42 |
| **HR (BPM)** |  | 69 ± 8 | 69 ± 7 | 0.96 |
| **Accelerometer (CPM)** |  | 36 ± 18 | 29 ± 13 | 0.25 |
| **REE (Kcal)** |  | 1479 ± 202 | 1418 ± 155 | 0.37 |
| **AEE (Kcal)** |  | 723 ± 270 | 710 ± 359 | 0.91 |
| **TEE (Kcal)** |  | 2447 ± 401 | 2364 ± 409 | 0.59 |

Body composition during the study period showed no detectable effect of the supplementation. The participants wore a combined accelerometer and heart rate monitor two days before and eight days after muscle injury to control for physical activity level in the critical period of the regenerating process. No difference between groups in input (beats per minute (BPM) and counts per minute (CPM)) or derivative parameters (resting energy expenditure (REE), activity energy expenditure (AEE) and total energy expenditure (TEE)) were detected (Table 1).

### Tolerability and compliance

In general NRPT supplementation was well tolerated. Two subjects reported obstipation in the placebo group whereas transient reflux (n=1) and transient loose stools (n=1) were reported in the NRPT group. The severity was mild in every case. In addition to low frequency of adverse reactions, compliance rates (measured by tablet counting at the end of the study) were 95.7% ± 4.9% and 95.8% ± 4.0% in the placebo group and the NRPT group, respectively. NRPT supplementation significantly increased whole blood levels of NAD^+^, NAAD, and Me2/4PY as well as Pterostilbene sulfate confirming compliance and uptake of the supplement (Figure 2).

**Fig. 2:**
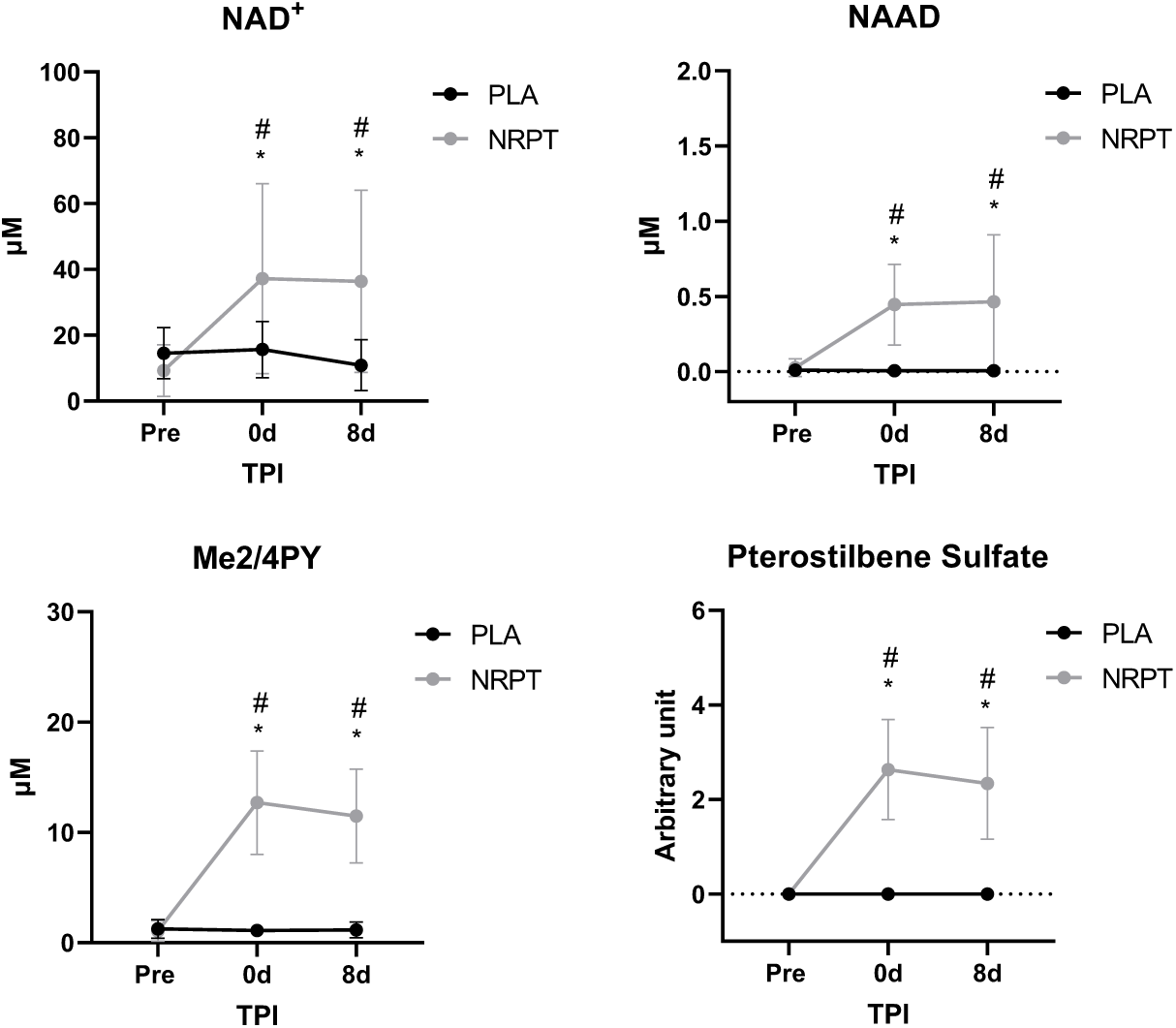
NRPT increases whole blood levels of NAD^+^ metabolites and Pterostilbene sulfate. Mass spectrometry analysis of whole blood revealed an increase in NAD^+^, NAAD, Me2/4PY, and Pterostilbene sulfate following NRPT supplementation. A level of steady state seemed to be reached before initiation of muscle injury. Data are expressed as mean ± SD. NRPT, n = 16; PLA, n = 15. # = p<0.05 between groups. *= p<0.05 vs. Pre. TPI = time post injury.

### NRPT does not improve muscle function after injury

The injury protocol induced significant clinical signs of muscle injury such as muscle soreness, swelling, decrease in muscle strength and release of muscle enzymes to the circulation (Figure 3). T2 signal from MR-imaging showed localized infiltration in the vastus lateralis part of the quadriceps femoris muscle 8 days post injury (Figure 3A). No difference in signal intensity was detected between NRPT and placebo. A pronounced elevation of p-myoglobin was observed already 2h post injury and remained elevated until 8 days post injury, whereas p-creatine kinase levels were only elevated 2 and 8 days post injury (Figure 3B+C). No supplementation effect was observed. Functional recovery, measured as maximal voluntary contraction (MVC) and rate of force development (RFD) of the quadriceps femoris muscle, were assessed at two knee angles (50 and 70 degrees). Both MVC and RFD were decreased after injury with no effect of the supplementation (Figure 3D). Accumulated muscle soreness (Area under the curve, AUC) indicated a tendency of lower soreness with NRPT supplementation (Figure 3E), however this difference did not reach statistical significance.

**Fig. 3:**
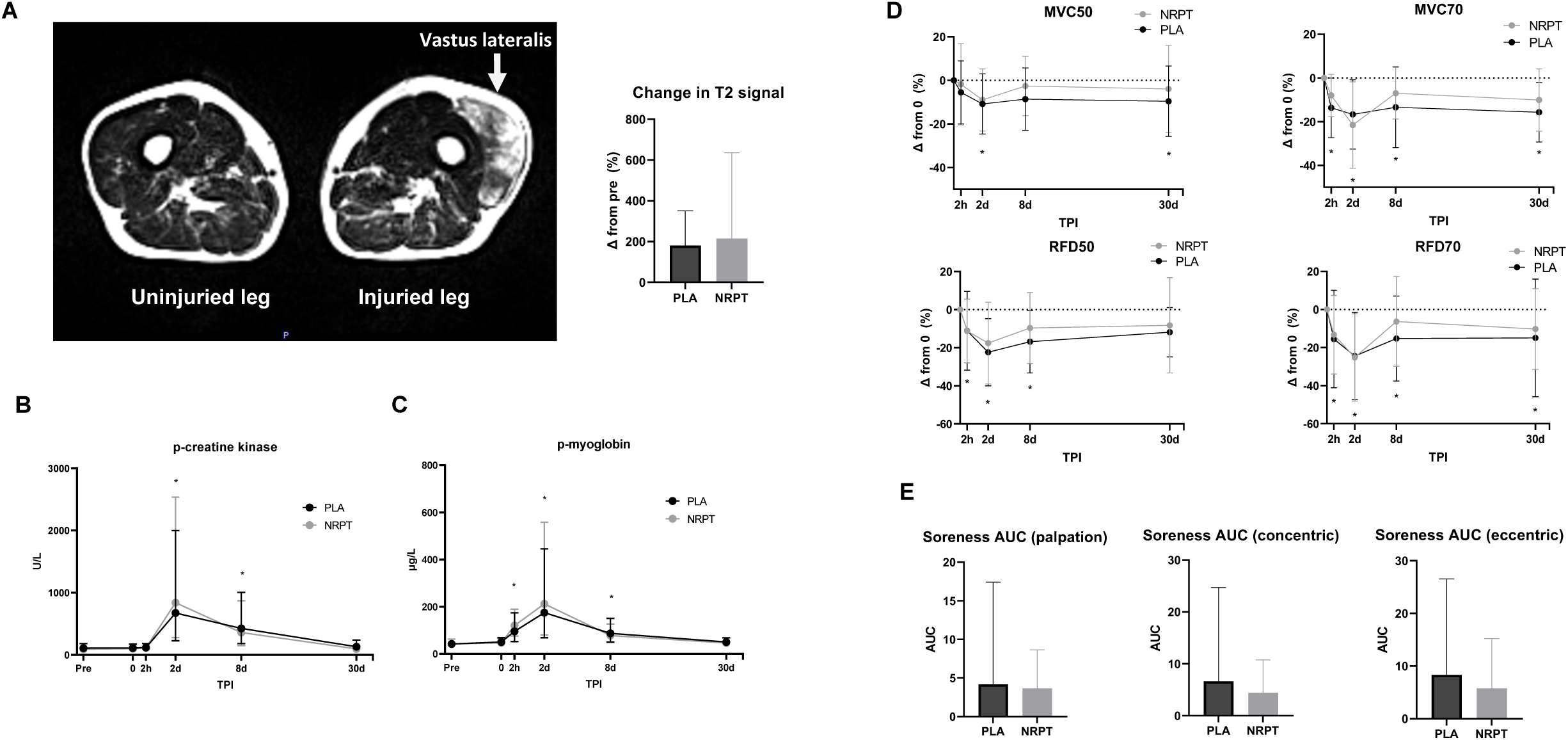
Clinical parameters associated with skeletal muscle injury. A: T2 weighted axial image of the thigh shows increased signal intensity in the vastus lateralis part of the quadriceps femoris muscle 8 days post injury but with no difference between groups (NRPT, n = 14; PLA, n = 12). Creatinine kinase (B), myoglobin (C), MVC + RFD (D), and muscle soreness (E) did response to injury but not to NRPT supplementation (NRPT, n = 16; PLA, n = 15). Data are expressed as geometric mean ± SD except from MVC and RFD which is expressed as mean ± SD. * = p < 0.05 vs. Pre. TPI = time post injury. MVC = maximal voluntary contraction. RFD = rate of force development. AUC = area under the curve.

### NRPT does not alter local cellular response to muscle injury

Flow-cytometry and Fluorescence-activated cell sorting (FACS) was utilized for quantification and sorting of MuSCs based on established protocols (22, 23). The MuSC content increased by 107% from pre injury to 8 days post injury with no difference between NRPT and placebo (p=0.58) (Figure 4A+B). To increase resolution of initial MuSC activation, cell size was measured by forward scatter from flow cytometry. 2 days post injury mean cell size was increased by 8% with a maximum increase of 18% observed 8 days post injury. No supplementation effect was detected (Figure 4C). To examine initial cell cycle entry of the MuSC pool, highly pure MuSCs were directly isolated and *ex vivo* EdU-incorporation was evaluated 48 h post sort. Pre injury, 23% of isolated MuSCs were positive for EdU which was increased by 113% and 96% 2 and 8 days post injury, respectively. This confirms a high proliferative rate of the MuSC pool at these two time points but with no effect of supplementation (Figure 4D+E). However, we did note a tendency towards a significant supplementation x time interaction in MuSC EdU-incorporation (p=0.05). Taken together, MuSC characteristics confirmed a large demand for recruitment of the MuSC pool in response to injury but did not reveal effects of NRPT supplementation.

**Fig. 4:**
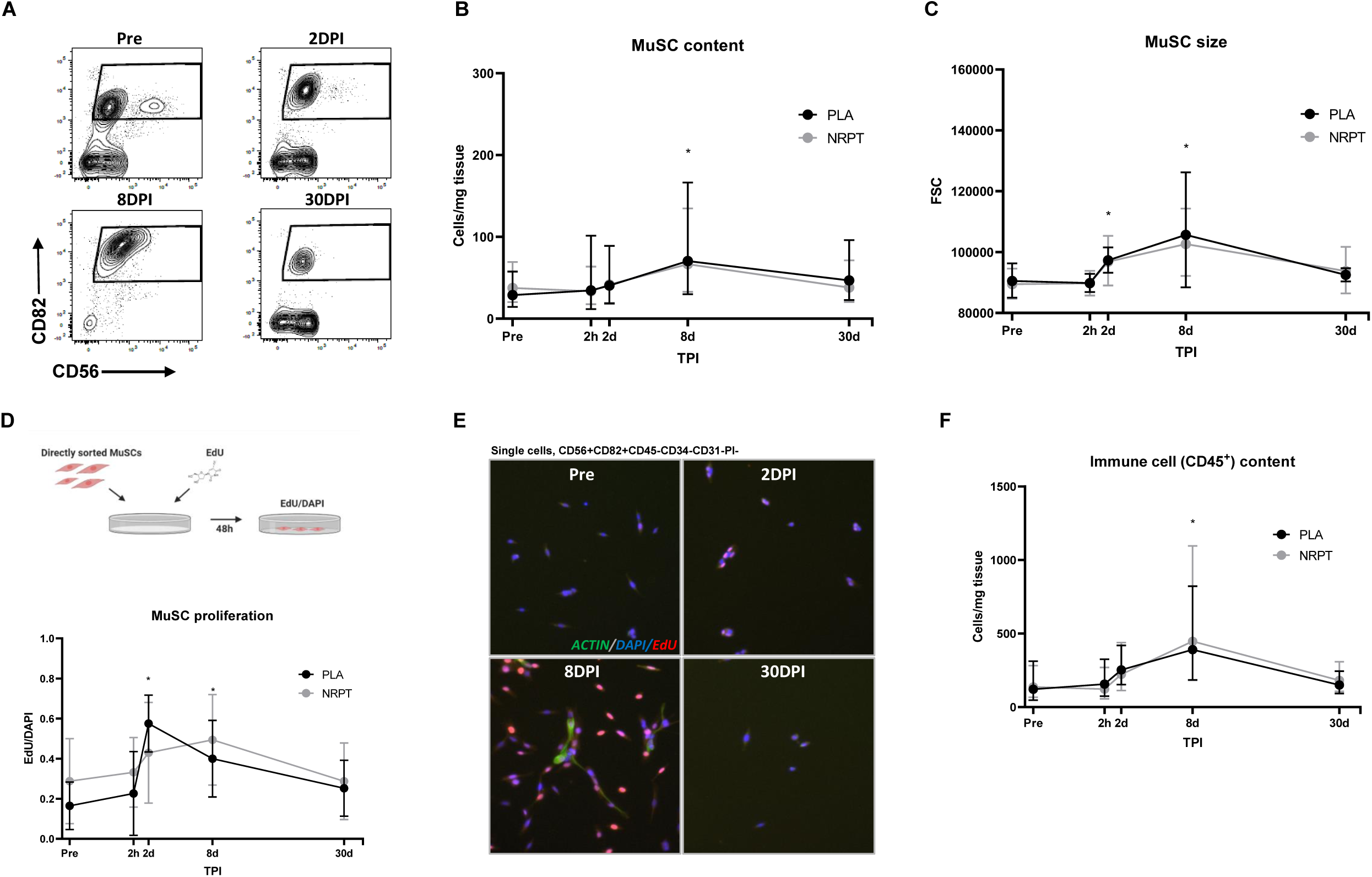
Muscle stem cell response to skeletal muscle injury. A: CD56 versus CD82 contour flow-plots of single CD45^-^CD31^-^CD34^-^PI^-^ cells from skeletal muscle biopsies Pre, 2, 8, and 30 days post injury. MuSCs number was quantified per milligram of skeletal muscle tissue (B) and size was measured from forward scatter (FSC) of flow cytometry (C). As a measure of MuSC proliferation, MuSCs were sorted and ex vivo EdU-incorporation was measured after 48h of incubation and expressed relatively to DAPI (D+E). F: Total content of hematopoietic cells (CD45^+^). Data are expressed as geometric mean ± SD except from MuSC proliferation which is expressed as mean ± SD. NRPT, n = 16; PLA, n = 15. * = p < 0.05 vs. Pre. DPI = days post injury. MuSC = muscle stem cell. TPI = time post injury.

The inflammatory response and its modulation is an essential part of the regeneration process and MuSC function, and we therefore examined the quantity of infiltrating hematopoietic cells (CD45^+^). Total number of CD45^+^ cells was increased by 236% 8 days post injury, but no effect of supplementation was observed (Figure 4F). Overall, these data show pronounced effects of the induced injury but with no effect of NRPT on cellular response to muscle damage.

### NRPT does not affect histological changes in response to injury

To study the effectiveness of the regenerative response, histological sections from biopsies were examined for changes in muscle fiber area and presence of newly generated fibers (internal nuclei and expression of embryonic Myosin Heavy Chain (eMHC)). As expected, mean fiber area was decreased 30 days post injury (Figure 5A). This was supported by left displacement of the fiber area distribution plot (Figure 5B) indicating an increase in small and newly generated fibers after injury. Muscle fiber area distribution was smaller in the NRPT group compared to the placebo group at all time points (including pre injury). The number of fibers containing internal nuclei and eMHC were increased 30 days post injury with no effect of NRPT supplementation (Figure 5C+D).

**Fig. 5:**
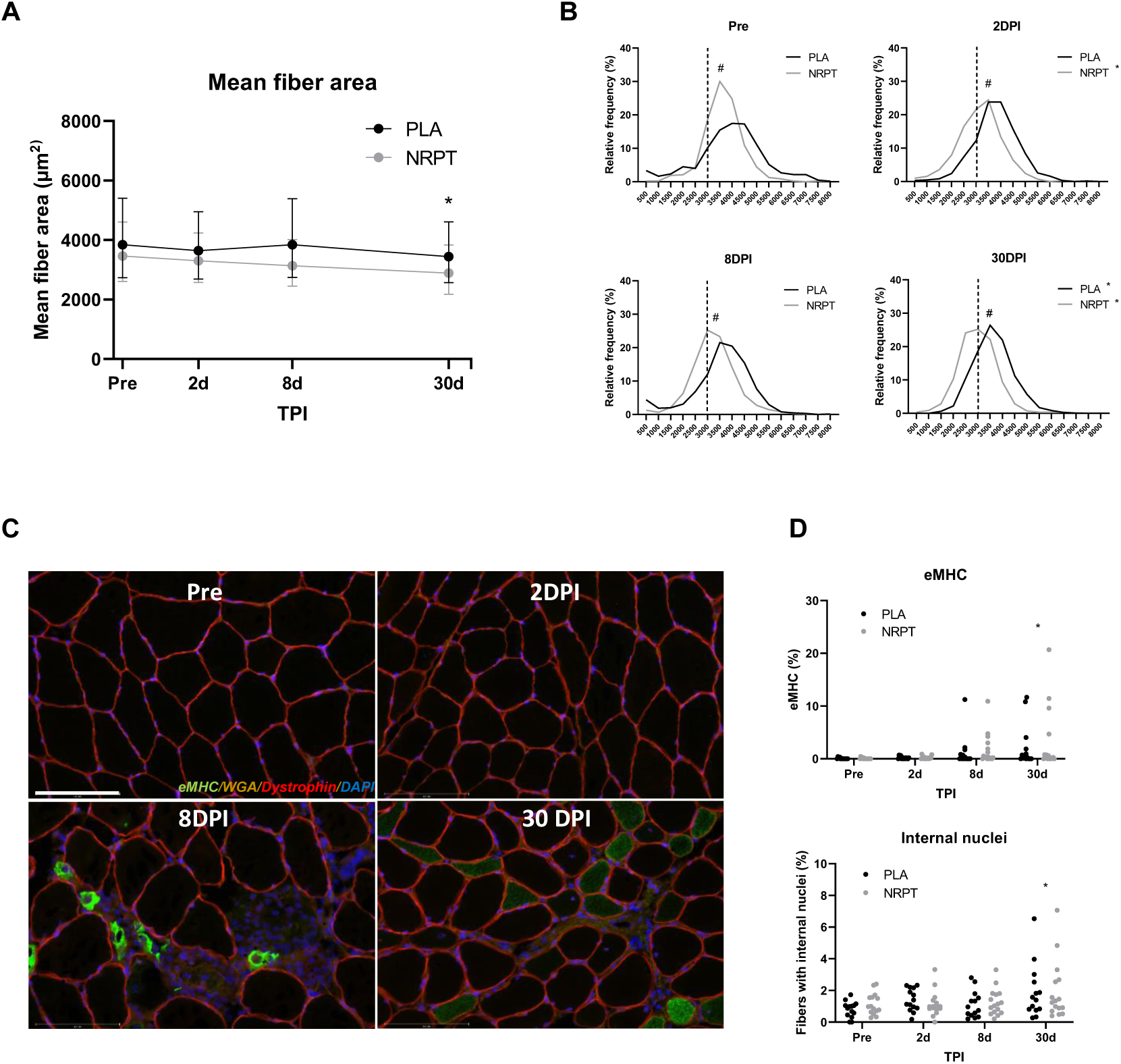
Histological analysis of myofibers. A: Mean fiber area was decreased 30 days post injury and this was supported by a left displacement of the fiber area distribution plot (B). Immunostaining revealed areas with infiltrated muscle fibers as well as clearly expression of embryonic myosin heavy chain (eMHC) (C). Both eMHC expressing fibers and number of internal nuclei were increased 30 days post injury (D). Data are expressed as mean ± SD. NRPT, n = 16; PLA, n = 15. # = p<0.05 between groups. *= p < 0.05 vs. Pre. TPI = time post injury. DPI = days post injury.

## Discussion

The main outcome of this study is that oral supplementation with NRPT does not improve recruitment of the MuSC pool after muscle injury in healthy, elderly subjects. The MuSC content 8 days post injury was chosen as primary endpoint in this trial. MuSC content is not a direct measurement of MuSC activation *per se*. However, the primary endpoint was supported by secondary outcomes such as MuSC size and cell-cycle entry kinetics (EdU-incorporation), and neither primary nor secondary endpoints supported a role of NRPT in treatment of muscle injury.

The injury protocol induced substantial skeletal muscle damage as confirmed by muscle edema (T2-signal on MRI), release of muscle enzymes to the circulation, reduced muscle function and soreness. By applying electric muscle stimulation protocol, we were able to induce more substantial skeletal muscle injury and subsequently regenerative response than injury protocols based on dedicated exercise or controlled voluntary eccentric muscle contraction. The quantities of muscle enzyme release in these elderly subjects were equivalent to application of a similar protocol in elderly men (60-73 years) but inferior to the response to injury in younger men (24) which probably reflects a greater quantity of muscle mass affected in the young. We found decreased muscle strength of the quadriceps femoris muscle by approximately 25% two days post injury. This underlines the potency of the protocol since T2 signal 8 days post injury documented that only the vastus lateralis part of the quadriceps femoris muscle was directly injured by the protocol. Furthermore, infiltration of immune cells and histological analysis of skeletal myofiber turnover (eMHC, internal nuclei and smaller muscle fibers after injury) confirms a substantial myofiber injury and subsequent regeneration up to 30 days after induction of injury which, to some extent, resembles that of experimental animal muscle injury protocols.

We found no effect of NRPT supplementation on either quantity or quality of MuSCs. In animals, both MuSC quality and quantity decline during ageing (3), and these declines have been shown to be partly reversible (16, 17, 25, 26). Human MuSC quantity also declines during ageing (27, 28), but the effect on MuSC quality is much less investigated. Recent evidence suggests that a major determinant of MuSC quality is the speed at which the cell can activate from quiescence to perform first cell division. Consequently, this speed of MuSC transition is essential for injury recovery rate in mice (29). In humans, the time to first cell division is shown to be approximately 83 hours for MuSCs (from the latissimus dorsi muscle) obtained from an uninjured muscle (30). To examine MuSC transition and cell cycle entry kinetics, we performed EdU-incorporation during the initial 48 h after isolation. This revealed a marked increase in cycling MuSCs at 2 days post injury, although the total content of MuSC was not yet increased. This was consistent with an increase in MuSC size 2 days post injury, suggesting that the MuSCs were indeed activated and undergoing an anabolic process in response to the injury which is similar to animal reports (29, 31). The increase in MuSC proliferation activity was maintained 8 days post injury while the MuSC size further increased. The latter is likely reflecting a larger pool of cycling MuSCs with a high metabolic and cellular turnover to support further cell divisions. Thus, 2 days post injury of MuSC size more readily represent cell cycle entry and cycling whereas 8 days and 30 days post injury may also reflect exit and committed myogenic differentiation. These data provide novel insight into the cell-cycle kinetics of human MuSCs in response to a muscle injury and suggest a high degree of similarity to data from animal models. Moreover, this approach brings forward a novel perspective on characterizing of mononuclear cells in skeletal muscle biopsies into human interventional studies.

Oral intake of NR can increase NAD^+^ levels in circulatating leucocytes as well as affect the skeletal muscle NAD^+^-metabolome in humans (21, 32-37). We found similar increases in NAD^+^ in whole blood which demonstrates compliance to supplementation, but this does not necessarily translate into uptake in human skeletal muscle and subcompartments such as MuSCs. Recent human trials have shown no effect of oral intake of NR on skeletal muscle mitochondrial content, structure, and bioenergitics (35-38) which contrasts findings in animals (17). This could be due to lower NR doses in human trials or because NR has been used in people with preserved NAD^+^ levels. However, proof of concept for improved muscle strength and mitochondrial biogenesis has been determined using nicotinic acid (NA), an alternative NAD^+^ precursor, in patients with Adult-Onset Mitochondrial Myopathy (39). These patients had both lower systemic and muscle NAD^+^ levels compared to healthy controls, which were rescued following four months of NA supplementation. Therefore, it is important not to preclude potential benefits of NRPT supplementation in populations suffering from low NAD^+^ levels.

Despite the randomized design, the mean age differed between the placebo group (63 years) and NRPT (69 years) group and this difference reached statistical significance. Therefore, a type two error of no supplementation effect cannot be excluded. However, we observed no correlation between age and MuSC response to injury, which could have explained the results (Figure S1). In addition, MuSC content, size, and proliferation did not correlate with age in pre injury biopsies (Figure S1). Finally, skeletal muscle fiber size is known to decrease during ageing, and we found a similar trend in the pre injury biopsy (data not shown). Thus, we cannot exclude that the greater frequency of small myofibers in the NRPT group is related to the difference in age. Overall, our findings of no effect of NRPT supplementation on recruitment of the MuSC pool does not seem to be driven by a difference in age.

While our study was not powered to determine effects of NRPT on all secondary endpoints we did notice a tendency towards a lower muscle soreness in NRPT group. Potential positive NRPT effects on muscle ache could be speculated to influence daily activity. However, physical activity and performance in muscle strength tests did not correlate with muscle soreness so indirect effects mediated by reduced muscle pain are unlikely. In the present study, we chose to determine the combined effect of NR and PT on muscle regeneration in elderly. The two previous human studies showing effects of NRPT on muscle strength used a duration of 2-4 month of supplementation (20, 21). An effect of longer duration of pre-injury supplementation, the individual components, or NRPT effects on skeletal muscle metabolism in uninjured conditions cannot be ruled out.

In conclusion, supplementation with NRPT at 1000/200 mg daily is safe but does not improve recruitment of the MuSC pool and thereby skeletal muscle regeneration after substantial injury in elderly human subjects.

## Material and methods

### Ethical approval

The study was conducted in accordance with the Declaration of Helsinki after approval by the local Research Ethics Committee in Central Denmark Region (1-10-72-301-18). The study was registered at clinicaltrials.gov (NCT03754842) before recruitment was commenced. Participants received oral and written information before written consent was obtained.

### Study population

32 men and women were recruited to the study through local media advertisement. Inclusion criteria were met, if: age: 55-80 yr (women should be postmenopausal), non-smokers, BMI: 20-28 kg/m2, <2 hours of physical activity/week and written informed consent. Exclusion criteria included endocrine disease, neurological or muscle disease, other severe disease or >30 min physical activity/day. Before enrollment, participants underwent a physical examination by a trained physician including routine clinical biochemistry and electrocardiography to evaluate eligibility for the study (Table 1). Three participants were treated for mild hypertension and received: Losartan, 50 mg (n=2) and Bendroflumethiazide + potassium chloride 2.5/573 mg (n=1). Four participants were using D-vitamin and calcium, three were using magnesium and zinc as a supplement. Participants were asked to refrain from supplementation with vitamins and dietary supplements two weeks before enrollment. 31 participants completed the study. One participant dropped out after the first visit as a result of pain related to the biopsy.

### Study design

The study was an investiga tor-initiated randomized, double-blinded, placebo-controlled trial in which participants received oral supplementation with Nicotinamide Riboside (NR) and Pterostilbene (PT) (NR: 500 mg, PT: 100 mg) (Basis™, Elysium Health, NY, USA) twice daily or placebo (capsules identical in number and external appearance). The Pharmacy at Aarhus University Hospital, Denmark was responsible for randomization, blinding, packaging, and labeling of the trial dietary supplement. Block randomization was used with a block size of four - two participants receiving placebo and two participants receiving NRPT in a random pattern. Participants and investigators were blinded to supplementation. Once all participants had completed the study, the randomization code was released. A detailed description of the study design can be seen in Figure 1.

### Muscle injury protocol

Participants were randomly assigned to injury in the dominant or non-dominant leg. Skeletal muscle injury was induced in adapted protocol from Crameri et al. (40) using electrically stimulated eccentric contractions. Participants were seated upright in a dynamometer (HumacNorm™ 1, CSMI Inc., Stoughton, MA, USA) and the chair position was adjusted so the center of the knee were aligned with the turning point of the dynamometer. The length of the arm was set three centimeter above the lateral malleolus. Stimulation patches (5 × 10 cm) were placed over the vastus lateralis muscle ∼10 cm below anterior superior iliac spine and 5 cm above patella. The stimulator (Elpha II/3000, Biofina, Odense, Denmark) delivered a current at a frequency of 35 Hz, 300 us pulse duration, 0-100 mA with an increasing phase of 0.5 sec, plateau of 5 sec and decreasing phase of 0.5 sec every 5^th^ sec. Applied mA were controlled by the participant, and they were regularly encouraged to go to highest tolerable level. When stimulation sufficiently caused extension in the knee joint to 20°, the lower leg was automatically pushed in the opposite direction by the dynamometer arm with a range of motion of 90°-20° (where 0° is full extension). 5 × 20 contractions were completed at 30°/second and 180°/second, respectively (200 contractions in total). Participants had a familiarization phase of 5-10 contractions without eccentric load before initiation of the protocol to obtain a sufficient stimulation level. Moment-angle curves were recorded from the contractions, expressed relatively to body weight, and total work calculated as sum of integrated curves using angles expressed in radians.

### Muscle strength test

Maximal voluntary contractions (MVC) and Rate of Force Development (RFD) of extension in the knee joint were measured in both legs in a dynamometer (HumacNorm™ 1, CSMI Inc., Stoughton, MA, USA) at two joint angels (70° and 50°). Three contractions were measured at each angel and lasted for 2-4 sec. with 30 sec. rest in between. Participants were encouraged to initiate the contraction as fast as possible. Muscle strength test were performed ∼20 min after muscle biopsy at all time points except from day 0. Here it was performed immediately after the muscle injury protocol. MVC was determined as the highest measured value. RFD was determined as the initial slope of the moment-time curve from 0-100 ms (with 0 as initiation of the contraction).

### Muscle soreness

Muscle soreness was assessed in both legs on a continuous visual analog scale ranging from 0-10 cm (0 = no pain, and 10 = worst thinkable pain) by; self-palpation of the vastus lateralis part of the quadriceps femoris muscle using three fingers (palpation), active flexion with no load (flexion), and squad (eccentric). Soreness was assessed as the first thing in the morning at pre, 0 (immediately before and after injury protocol), 1, 2, 3, 4, 5, 6, and 7 days post injury. Participants received supervision the first three times. Area under the curve (AUC) of soreness score (y-axis) and time (x-axis) was compared between groups.

### Blood analysis

Blood analysis of creatinine kinase, myoglobin and leucocytes were carried out at the Department of Clinical Biochemistry, Aarhus University Hospital. Whole blood for NAD^+^ metabolites and Pterostilbene sulfate analysis were collected in EDTA tubes and immediately placed on ice until storage at -80°C.

### UHPLC-MS analysis

Method was developed according to Lu et al. (41) and was modified in order to be used for the specific matrix and instrumentation. Whole blood samples were blinded and randomized before analysis. 100 µl of whole blood was mixed with 20 µl of isotopically labeled internal standard mix and 380 µl of extraction solvent (methanol: water = 8: 2; V: V). Internal standard mix included 10 mg/l of: nicotinic-d4 acid, nicotinamide-d4, N-methylnicotinamide-d4, D-tryptophan-d5 (all from CDN isotopes, Quebec, Canada), adenosine-^13^C_5_ and nicotinamide adenine dinucleotide-d4 (Toronto Research Chemicals, Toronto, Canada) and adenosine-^13^C_10_,^15^N_5_ 5′-triphosphate (Sigma Aldrich, Darmstadt, Germany). All internal standards were dissolved in methanol and were used to control extraction efficiency and method variation. Extraction was made using ultrasound ice cold bath for 10 minutes. Proteins were removed overnight at -20°C. Before centrifugation (10 000 rpm, 5 min, 4 °C), samples were re-equilibrated at the room temperature. After centrifugation, 180 µl of supernatant was taken and evaporated to dryness under a stream of nitrogen and reconstituted with 50 µL of methanol: water = 8: 2 (V: V). Calibration curves were prepared in the same way by replacing sample with metabolite standards.

NAD^+^ metabolites and Pterostilbene sulphate were analyzed using ultra-high performance liquid chromatography with mass spectrometry detection (UHPLC-MS). Liquid chromatograph (Agilent, 1290 Infinity II, Santa Clara, CA, US) was coupled with mass spectrometer (timsTOF Pro, Bruker Daltonics, Bremen, Germany). Metabolite separation was achieved using a 1.7 µm, 2.1 × 150 mm Waters ACQUITY Premier BEH Amide column (Waters, Milford, MA, USA). Mobile phase A was miliQ water including 10mM ammonium acetate and 5µM of medronic acid. Mobile phase B was composed of acetonitrile and water (V: V = 9: 1) again with the same amount of ammonium acetate and medronic acid. Gradient started with 90 % of mobile phase B where it was stable for 2 min. Thereafter, it was decreased to 55% in 10 min where it was hold for 2 min. At the end, the initial composition of mobile phases was restored. Injection volume was 2 µl. Mass detector was set to analyze exact mass spectra in full scan mode in the range from 50 to 1000 Da, at 1Hz. Mass resolution was >60,000. Metabolites were identified by exact mass and retention time of metabolite standards with exception of pterostilbene sulfate, which was confirmed using fragmentation spectra. Quantification was made with dilution series of metabolite standards. Concentration was expressed in µM of metabolite in blood. Pterostilbene sulfate was expressed as Pterostilbene equivalent.

### Dual-energy X-ray Absorptiometry

Body composition were assessed by a whole-body DXA scan at Department of Endocrinology, Aarhus University Hospital (Hologic Discovery, Hologic). Two participants in the NRPT group did not underwent the second scan.

### MR-imaging

MR-imaging was carried out pre and 8 days post injury on a Siemens Skyra 3T MR scanner (Siemens, Erlangen, Germany) at the MRI Research Centre, Aarhus University. Participants were placed supine on the scanner bed and an 18 element anterior coil was positioned over both thighs. Participants were carefully instructed not to move the lower extremities during imaging. Localizer images were acquired to define the length of the vastus lateralis muscle of the quadriceps femoris. The amount of edema in the muscles was assessed by T2-maps, which were acquired by a multi-echo spin echo sequence with Field-of-View of 420 × 289 mm, matrix of 256 × 176 and 8 slices with thickness of 7 mm. Repetition time was 2.7 s and 16 echoes with echo times from 20 ms to 320 ms were acquired. Scan time was 3 minutes. T2-maps were obtained by fitting an exponential function to the images.

T2 signal was quantified in PMOD version 4.0 (PMOD Technologies Ltd, Zurich, Switzerland). A volume of interest was generated as 3-4 axial images in the middle of the vastus lateralis muscle of the quadriceps femoris, where the muscle is well-defined. Due to technical issues, data from only 26 individuals were obtained.

### Activity level and energy expenditure

To control for activity level and energy expenditure, participants wore a combined heart rate monitor and accelerometer (Actiheart 4, CamNtech Ltd., Cambridgeshire, United Kingdom) horizontally attached over the left third intercostal space using electrocardiogram electrodes (cat. no.: SP-50, Pulse Medical, United Kingdom). The Actiheart was initialized using 60 sec epochs and synchronized with a digital clock before mounting. The software provides an estimate of energy expenditure using branched modelling considering both HR and activity measurements with input of resting HR, weight and age. Data on, activity (counts per minute (CPM)) and mean HR (beats per minute (BPM), resting energy expenditure (REE), activity energy expenditure (AEE), total energy expenditure (TEE) were extracted from the Actiheart software. A detailed software/model description can be found elsewhere (www.camntech.com). The device was worn from 2 days pre injury to 8 days post injury. Days with > 120 lost epochs were excluded from analysis. Data collection was lost in one participant from the placebo group and one participant in the NRPT group. Overall, an average of 7.9 days per participant were included in the analysis.

### Muscle biopsy

Muscle biopsies were obtained from the vastus lateralis muscle with a Bergström needle with manual suction under local analgesics (Xylocain 10 mg/ml, AstraZeneca, Stockholm, Sweden) in sterile conditions. Pre injury biopsy was collected from the non-injured leg and four biopsies were collected from the injured leg (2h, 2, 8 and 30 days post injury). To minimize the effect of repeated sampling, a minimum distance of 3 cm between incisions was ensured. In addition, participants were stratified into three different biopsy location patterns. Biopsy material adequate for histological analysis was parallel aligned and embedded in Tissue-Tek (Sakura Finetek Europe, Zoeterwoude, The Netherlands) and frozen in precooled isopenthane and stored at -80°C. Biopsy material for Fluorescence Activated Cell Sorting (FACS) was weighted and stored in C-tubes (cat. no.: 130-093-237, Miltenyi Biotec, Lund, Sweden) containing 6.5 ml ice-cold wash buffer [(Hams F10 incl. glutamine and bicarbonate) (cat. no.: N6908 Sigma, Sigma-Aldrich, Denmark), 10% Horse serum (cat. no.: 26050088, Gibco, ThermoFisher Scientific, MA, USA), 1% Penstrep (cat. no.: 15140122, Gibco)] for a maximum of 2h before digestion.

### Tissue digestion

Prior to digestion of muscle tissue, Collagenase II (cat. no.: 46D16552, Worthington, Lakewood, NJ, USA) and Dispase II (cat. no.: 04 942 078 001, Roche Diagnostics, Basel, Switzerland) were added to the C-tube making a final concentration of 700 U/ml and 3.27 U/ml, respectively. Mechanical and enzymatic muscle digestion was then performed on gentleMACS (cat. no.: 130-096-427, Miltenyi Biotec) for 60 min using a skeletal muscle digestion program (37C_mr_SMDK1). The solution was added 10 ml wash buffer and filtered through a 70µm cell strainer and washed twice to collect any remaining cells before centrifuged at 500g for 5 min. The supernatant was removed and cell pellet resuspended in freezing buffer (cat. no.: 130-109-558, StemMACS, Miltenyi Biotec) and stored at - 80°C.

### Flow cytometry and cell isolation

The protocol and antibody panel for flow cytometry and cell isolation has been validated and described in details elsewhere (22). Cells were sorted into 5 ml collection tubes containing 500µl wash buffer at 4°C. Gating strategies were optimized using Fluorescence Minus One (FMO) controls for each antibody. Data was collected in FACSdiva software and analyzed in FlowJo™ Software Version 10.6.2.

### Cell proliferation

To detect cellular ex vivo proliferation, we utilized 5-ethynyl-2’-deoxyuridine (EdU)/click-it assay visualizing cells containing newly synthesized DNA (cat. no.: C10337 and C10340, Invitrogen). Cell population bulk experiments were carried out in 96-well half-area plates (Corning^®^, NY, USA) coated with ECM (cat. no.: E1270, Sigma). Cells were plated in wash buffer + 10 µM EdU immediately after sorting. After 48h, cells were fixed in 4% paraformaldehyde for 8 min and washed 5 min x 3 in PBS and kept in PBS at 4°C until further analysis. To detect EdU-incorporation, we followed the manufacturer instructions and counter-stained the cells with DAPI (1:50.000, cat. no.: D3571, Invitrogen, Thermo Fisher Scientific). Images were acquired using an EVOS M7000 automated imaging system (Thermo Fisher Scientific) and performed automatically to ensure similar treatment of all wells. EdU- and DAPI-positive cells were semi-automatically counted in ImageJ and expressed relative to each other.

### Histology and immunohistochemistry

Cryosections of biopsies form pre, 2, 8, and 30 days post injury were used for histology (thickness of 10 µm). After thawing to room temperature, sections for immunohistochemistry were fixed for 5 min in Histofix (cat. no.: 01000, Histolab Products AB, Västra Frölunda, Sweden) followed by blockade using 1% BSA + 10% FBS + 0.5% Triton in 1X PBS for 60 min at room temperature. Primary antibodies targeting eMHC (mouse, 1:8, F1.652, Developmental Studies Hybridoma Bank (DSHB), IA, USA) and Dystrophin (rabbit, 1:500, cat. no.: ab15277, Abcam, MA, USA) in 1% BSA + 10% FBS in 1X PBS were added and incubated overnight at 4°C. Sections were then washed 2×5 min in PBS and incubated with secondary antibody Alexa-fluor 488 goat-anti-mouse and Alexa-fluor 647 goat-anti-rabbit (1:500, cat. no.: A-11001 and cat. no.: A-21245, Thermo Fisher Scientific) combined with Wheat Germ Agglutinin (WGA) Texas Red™-X Conjugate (1:500, cat. no.: W21405, Thermo Fisher Scientific) for 60 min at room temperature. Finally, sections were washed in PBS 3×5 min, with one wash containing DAPI (1:50.000, cat. no.: D3571, Invitrogen, Thermo Fisher Scientific), and then mounted with cover slides. Minus primary controls were included for optimization to ensure specificity.

### Imaging and quantification

Images were acquired using EVOS M7000 automated imaging system (Thermo Fisher Scientific). For embryonic Myosin Heavy Chain (eMHC), internal nuclei, and fiber area quantification whole biopsy sections were scanned and stitched to one image. Areas containing length cut fibers were removed from analysis. eMHC and internal nuclei were manually counted in ImageJ and expressed relative to total amount of fibers in given section. An average of 527±188 were analyzed per biopsy.

For fiber area quantification Semi-automatic Muscle Analysis using Segmentation of Histology (SMASH) (42) were used on dystrophin stained sections. An average of 389±126 fibers were analyzed per biopsy.

### Statistics

Data are expressed as mean ± standard deviation if normal distributed or geometric mean ± 95% if not normal distributed. Data distribution was evaluated by inspection of QQ-plots. A Student’s t-test was performed to compare anthropometric, work, current, activity soreness and MRI data. P-Myoglobin (log-transformed), p-creatinine kinase (log-transformed), MVC data, MuSC data (log-transformed), CD45^+^ data (log-transformed), mean fiber area (log-transformed) were analyzed using a repeated-measurement mixed model analysis with intervention (placebo/NRPT), time, and the interaction between intervention and time as factors. In case of a significant interaction, pairwise comparisons were performed for differences within and between interventions and time and Dunnett’s method of multiple comparisons was performed. Chi-squared test was used to evaluate distribution of skeletal muscle fiber area. For internal nuclei and eMHC each group was tested separately with the Friedman test and with Dunnett’s method for multiple comparisons if the outcome was significant. To test for a supplementation effect, groups were compared at each time points post injury with the Mann-Whitney test. Significance was set at p<0.05. All analyses were carried out using GraphPad Prism version 8.3.0 (GraphPad Software) and STATA version 16. Graphs were generated in GraphPad Prism and figures were created with Biorender.com.

## Data Availability

All data are available upon request.

## Acknowledgements

We thank FACS core facility at Aarhus University, Denmark for technical assistance related to FACS and flow-cytometry. The authors would also like to thanks Lenette E. Pedersen and Pia B. Hornbek, Medical Research Laboratory, Department of Clinical Medicine, Aarhus University, Denmark for excellent technical assistance. Furthermore, we thank Elysium Health for providing active and matching placebo pills. Finally, we would like to thank all participants for their engagement to the study.

## Authors’ contributions

JBJ, JF, OLD, ABM, SR, NM, JTT, and NJ conceived and designed the experiments. JBJ, JF, TBB, SR, and KT performed the experiments. JBJ, JF, OLD, ABM, RWD, SR, ED, SC, KT, TM, JTT, and NJ analyzed and interpreted the data. JBJ and JF prepared the figures and drafted the manuscript. All authors read, revised, and approved the final manuscript. NJ is the guarantor of the work and has full access to all the data in the study, and takes responsibility for the integrity of the data and the accuracy of the data analysis.

