## Supplemental fig. S1 for "A randomized placebo-controlled clinical trial of Nicotinamide Riboside and Pterostilbene supplementation in experimental muscle injury in elderly subjects"

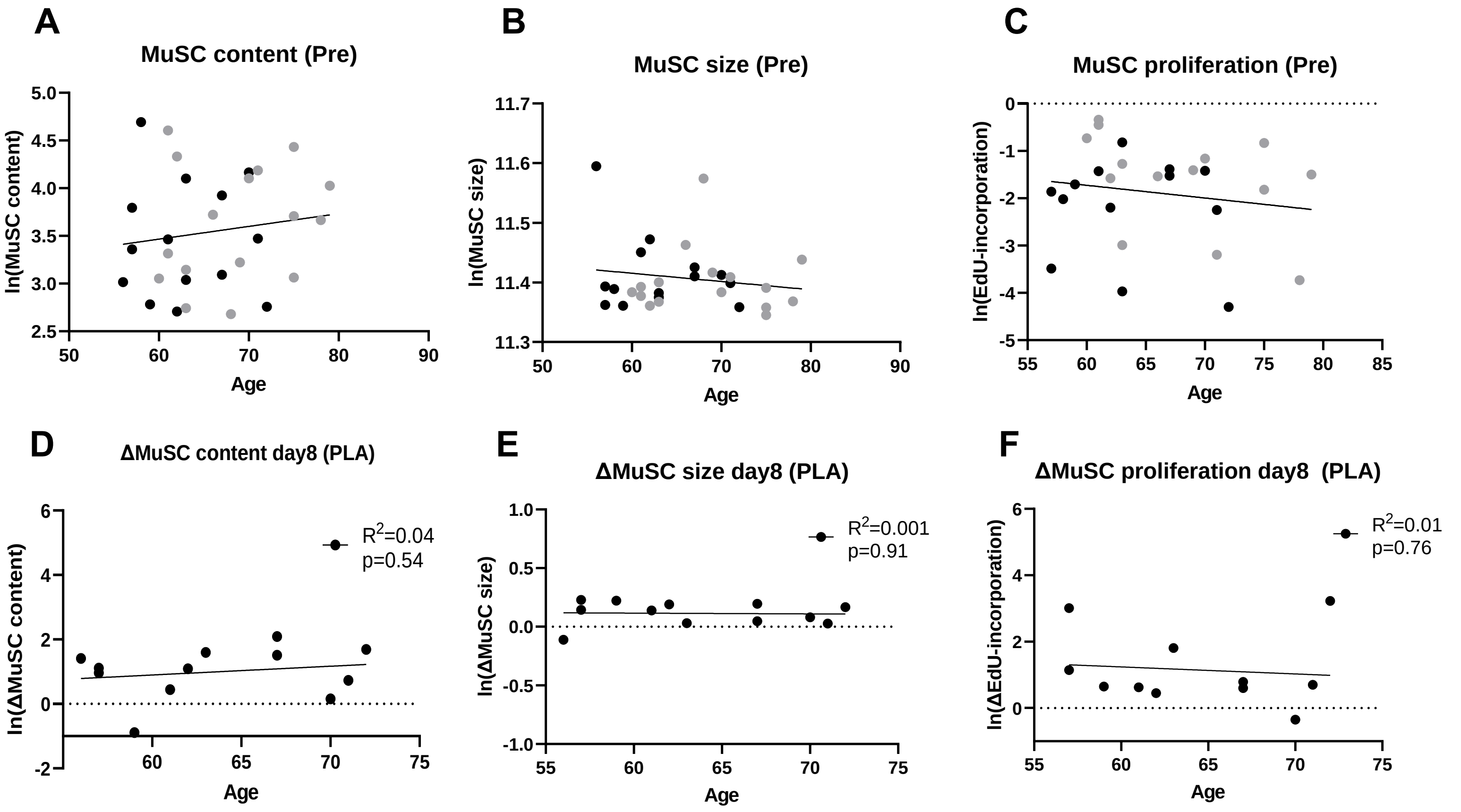

A
MuSC content (Pre)
ln(MuSC content)
Age
B
MuSC size (Pre)
ln(MuSC size)
Age
C
MuSC proliferation (Pre)
ln(EdU-incorporation)
Age
D
ΔMuSC content day8 (PLA)
R2=0.04
p=0.54
ln(ΔMuSC content)
Age
E
ΔMuSC size day8 (PLA)
R2=0.001
p=0.91
ln(ΔMuSC size)
Age
F
ΔMuSC proliferation day8 (PLA)
R2=0.01
p=0.76
ln(ΔEdU-incorporation)
Age

***S1: Correlation between MuSC parameters and age***

Correlation between age and MuSC content (A), size (B), and EdU-incorporation (C) from Pre biopsy. Correlation between age and change in MuSC number (D), size (E), and EdU-incorporation (F) from Pre to 8 days post injury in the placebo group.
